# Patient digital twins: an introduction based on a scoping review

**DOI:** 10.1101/2024.02.20.24303096

**Authors:** David Drummond, Apolline Gonsard

**Author notes:** Corresponding author: David Drummond, PhD, Department of Paediatric Pulmonology and Allergology, University Hospital Necker-Enfants Malades, AP-HP, 149 rue de Sèvres 75015, Paris, France.

## Abstract

The concept of digital twins, widely adopted in industry, is entering healthcare. In this scoping review, we analysed definitions and characteristics of patient digital twins being developed for clinical use. Searching for studies claiming digital twin development/evaluation until August 2023, we identified 86 articles representing 80 unique claimed digital twins, nearly all (98%) in preclinical phases. From the analysis of definitions and characteristics, we propose to define patient digital twin as “a viewable digital replica of a patient, organ, or biological system that contains multidimensional, patient-specific information”. Two main forms were found: simulation digital twins using computational modelling of patient anatomy/physiology to run personalised outcome predictions and therapy evaluations, mostly for one-time assessments; and monitoring digital twins harnessing aggregated patient data for continuous risk/outcome forecasting over time and care optimisation. As patient digital twins rapidly emerge, the proposed definitions and subtypes offer a framework to guide research into realising the potential of these personalised, integrative technologies to advance clinical care.

## INTRODUCTION

Each industrial revolution has transformed the practice of medicine. The first two led to the development of new techniques for the industrial collection of new data (biological, imaging, etc.) on the human body. The third - or digital - revolution transformed this analogue data into digital data, accelerating the exchange of information, and allowing the emergence of computer models to propose a diagnosis, establish a prognosis, and recommend a treatment.^1^

For some contemporaries, we are currently in the midst of the fourth industrial revolution, which is the merging of the physical and digital worlds, based on three pillars: the Internet of Things, increasing connectivity, and machine learning-based decisions.^2,3^

Digital twins are an emblematic illustration of the applications of this new industrial revolution. Michael Grieves introduced the concept in 2002 as a system consisting of a physical product, its virtual counterpart and two-way data exchange between the two entities.^4^ However, the term “digital twin” was first used and defined in 2010 by NASA engineers as “an integrated multi-physics, multi-scale, probabilistic simulation of a vehicle or system that uses the best available physical models, sensor updates, fleet history, etc., to mirror the life of its flying twin”.^5^ Following the popularisation of the concept of the fourth industrial revolution in 2015, digital twins attracted interest from all industries.^3^ With the objective of reducing production times through monitoring, coordination and control of production systems, the manufacturing industry became the most active sector in terms of research and implementation of digital twins.^6^ Digital twins also extended to construction, energy, transport, smart cities, agriculture, education and health.^6^

In the health sector, the concept of digital twins attracted interest from industry, scientists, clinicians and patients.^7,8^ Besides digital twins of hospitals, there is a strong rationale for the development of digital twins of patients, as these systems could offer personalised medicine through information gathered by the Internet of Things, real-time adaptation of treatments through efficient connectivity, and even automation of certain aspects of medical management through predictions based on machine learning.^9^ An increasing number of scientific publications claim to be developing or to have developed “digital twins” of organs, physiological systems, or patients. However, when the term “digital twin” is used in this context, it remains unclear if it corresponds to the definitions used in industry, or whether unique concepts and characteristics emerge for patient digital twins.

The three previous systematic literature reviews related to digital twins in health did not focus on patient digital twins.^10–12^ To contribute to the understanding of patient digital twins, we conducted a scoping review to systematically map research in this area. The following research question was formulated: “What are the definitions and characteristics of digital twins provided by research teams developing patient digital twins for clinical applications, as reported in the scientific literature?”

## METHODS

We conducted this scoping review in accordance with the Joanna Briggs Institute (JBI) guidelines for scoping reviews^13,14^ and reported it following the Scoping Review Extension of the Preferred Reporting Items for Systematic Reviews and Meta-Analyses (PRISMA-ScR) checklist.^15^ The authors received training on the methodology of scoping reviews using the JBI Reviewers’ Manual 2020.^16^ We registered the protocol for this review at the Open Science Framework (https://osf.io/7twn8/).

### Search Strategy

We created search strategies with a medical librarian to identify published articles on patient digital twins. We initially searched MEDLINE via PubMed. The text words contained in the titles and abstracts of relevant articles and the MeSH terms describing those articles informed development of a full search strategy. We adapted this search strategy for each included database and information source, namely: PubMed, Scopus, Embase, IEEE Web of Science, and Google Scholar. When searching Google Scholar, we used incognito mode to minimize effects of past search histories and screened the first 500 results. Full search strategy is reported in the appendix (pp3-4).

### Selection process

We collated the identified citations from the comprehensive search and uploaded them into Rayyan, an online review tool.^17^ Two independent reviewers (DD, AG) removed duplicates and screened titles and abstracts for inclusion based on the predefined criteria described below. All screening was performed in a masked, duplicate fashion. Any disagreements between reviewers were resolved through discussion. Reasons for exclusion at the stage of full-text screening were recorded.

### Inclusion and exclusion criteria

We included peer-reviewed research articles in which the authors claimed to be developing or to have developed/tested a digital twin in healthcare, subsequently referred to as a ‘claimed digital twin’ (CDT). Only literature published in English, French, or German was included up to August 31 2023. Finally, to match the context of this review, we restricted included research to articles focused on developing or testing digital twins specifically representing patients, or components of patients.

Exclusion criteria were: (1) animal studies, abstracts only, conference papers, reviews, editorials or correspondence, non-peer-reviewed articles; (2) studies in languages other than English, French, or German; (3) studies unrelated to health; (4) digital twins not representing patients or part of a patient; and (5) studies limited to proposing a framework for a digital twin with no case study.

### Data charting

Two reviewers (DD, AG) jointly developed a data charting form using Microsoft Excel to determine which variables to extract. For each included study, we extracted the following characteristics: study title, publication year, first author name, country of the corresponding author, and journal. We compiled the definitions of digital twins provided by the study authors and summarised their various dimensions (e.g. digital replica, high-fidelity representation, two-way data exchange). Drawing from NASA’s digital twin definition, we assessed whether the developed digital twins were multi-scale, integrated multi-physics modelling, and multiple data sources. We incorporated additional descriptors specific to digital twins for patient care, including medical discipline, organs/systems represented, categorisation of whether the models constituted anatomical representations (e.g. 3D) and/or models of physiological systems, and the study’s objective (simulation, prediction, monitoring, visualisation, generation of synthetic patients). Technical digital twin characteristics extracted included: types of patient data used, approach used (mechanistic, data-driven, hybrid), whether the digital twin included analytics and advanced visualisation, constituted a model for simulations versus a simulation itself, capacity and frequency of updates, inclusion of none, one-way or two-way data exchange with the patient, and for two-way exchange, the nature of feedback (recommendation or other type). Finally, we determined the clinical research phase of each digital twin by deriving the clinical research phases developed for artificial intelligence in medicine.^18^ Additional information on each variable is presented in appendix (pp 5-6).

To ensure consistency, two reviewers (AG, DD) independently extracted data from the first 10 articles prior to full data extraction. Any disagreements were resolved through discussion. Following this pilot extraction and adaptation of the data charting form, we extracted data from the remaining studies in a masked, duplicate manner. Studies not meeting inclusion criteria were excluded. Any disagreements between reviewers were resolved through discussion.

### Unsupervised classification of claimed digital twins

To perform unsupervised classification of CDT, we analysed dissimilarities by employing the Gower distance metric on a subpart of our dataset (the variables included are marked with an asterisk in appendix 2 (pp 5-6)).^19^ The optimal cluster count was identified using the Partitioning Around Medoids (PAM) method, guided by silhouette width analysis across 2 to 10 clusters.^20^ We then employed t-Distributed Stochastic Neighbor Embedding (t-SNE) for dimensionality reduction to facilitate visualisation in a 2D space. These analyses were performed in R (v.4.2.3) with the packages cluster, Rtsne, and ggplot2. We finally compared the characteristics of the CDTs of each cluster using the package gtsummary.

## RESULTS

### Characteristics of included studies

A total of 7,224 citations were identified from the search strategies (Figure 1). After removing duplicates and screening titles and abstracts, 154 articles were examined in full-text, and 68 were excluded (see appendix pp 7-11 for a full list of excluded full text reviews with exclusion reasons). Finally, 86 articles^21–106^ representing 80 unique CDTs were included in this scoping review. The full results, with individual data for each study, can be viewed in tabular form at the following address: https://osf.io/7twn8/.

**Figure 1:**
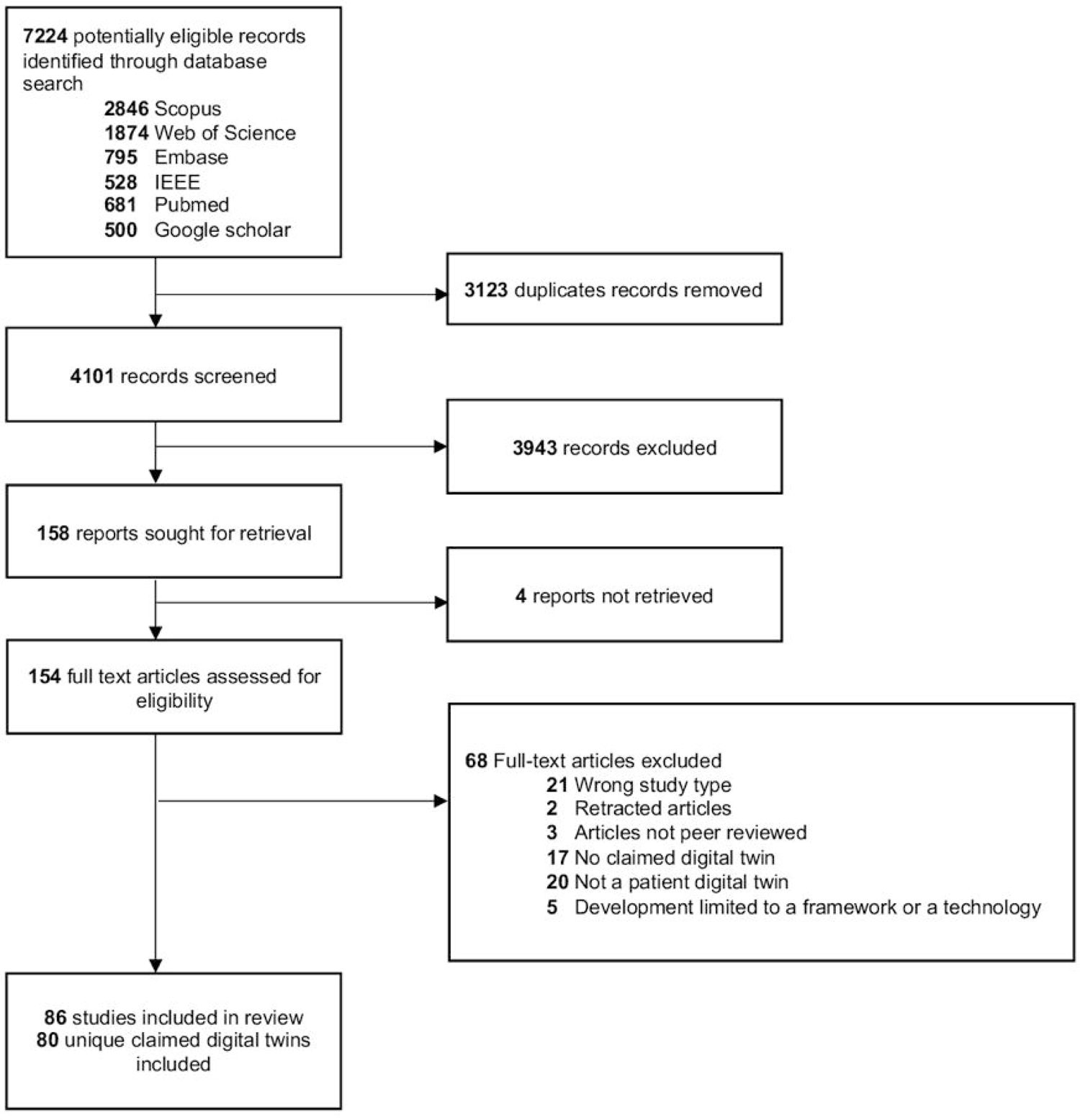
PRISMA flowchart

The first publications of CDTs appeared in 2019. Since then, publication volume increased yearly from 3 papers in 2019 rising to 23 in 2022 and 32 papers in the partial 2023 year (appendix p 12). Though incomplete, 2023 totals through August 31 still reflect rising publication volume.

The included publications originated from 22 countries (appendix p 12). The United States of America was the most common country of origin, with 22 articles (25% of included publications). At the continental level, Europe was leading with 43 publications (49%), followed by North America with 25 (29%), and Asia with 16 (18%). No articles from South America or Africa were found.

### Definitions of digital twin provided by authors

Definitions of “digital twin” were provided in 55 articles (63% of publications, appendix pp 13-16). Among articles with definitions, a digital twin was defined as a digital replica of a real object in 42 publications (76%), with real-time update in 23 (42%), a patient-specific approach in 13 (24%), and a two-way communication between the real and the digital object in 8 (15%) (Figure 2).

**Figure 2:**
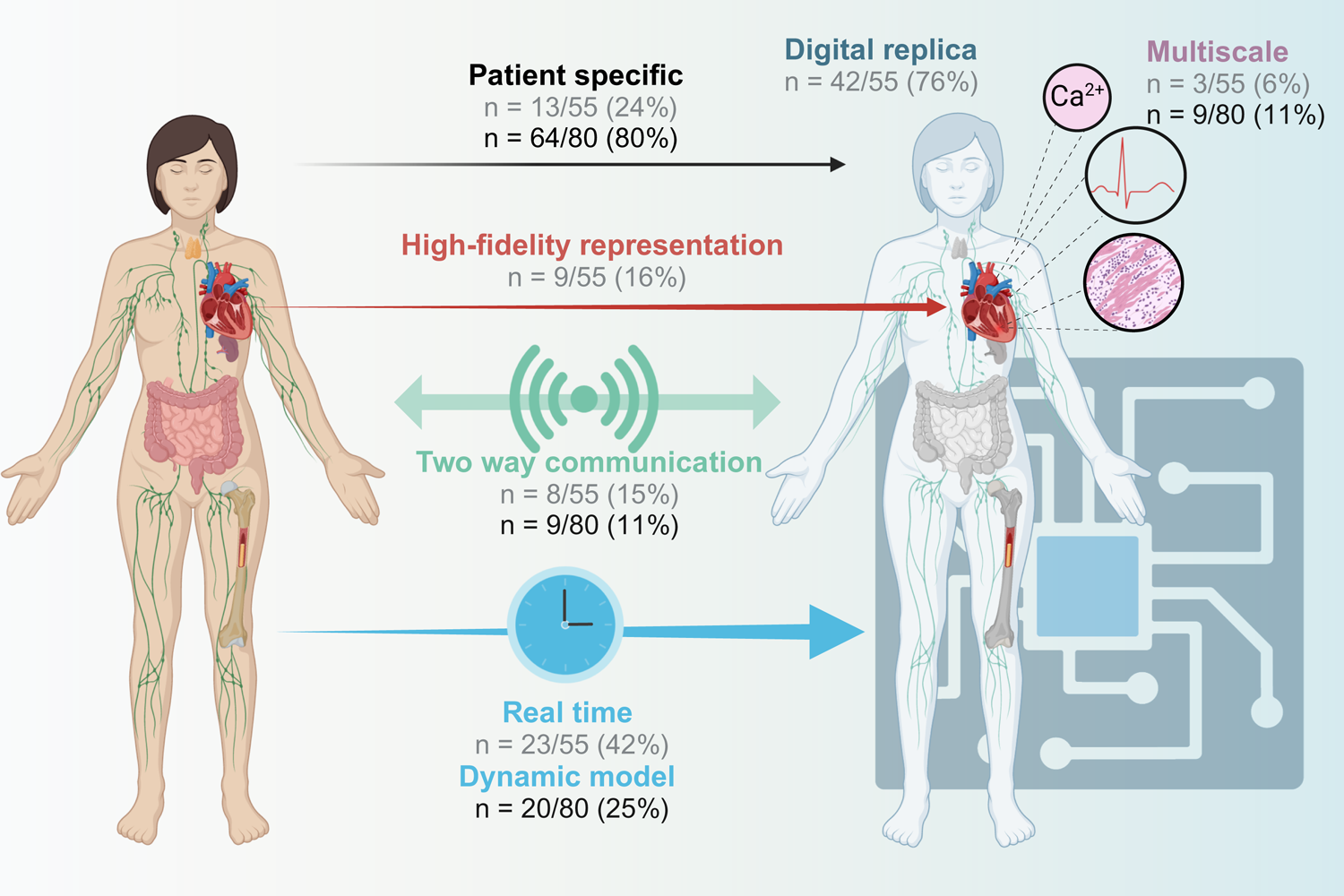
Dimensions of digital twins included in definitions provided by authors (grey) and included in the claimed digital twins (black). Created with Biorender.

### Medical domains

Of the 80 CDT identified, 48 (60%) represented specific organs or anatomical regions, while 11 (14%) embodied biological systems (e.g. immune system). The remaining 21 (26%) publications described other types of CDT (e.g. systems for emotion recognition, foetal heart monitoring, etc.). Among the 48 publications with CDT representing specific organs, the most widely modelled organs were the heart (15 CDT, 31%), the bones and joints (10, 21%), the lung (6, 12%), and the arteries (5, 10%) (Figure 3 and appendix p 17). Three (6%) CDT involved multiple organs. Among the 11 CDT representing biological systems, the endocrine (4, 36%) and the immune system (3, 27%) were the most widely involved (appendix p 17). The most highly represented medical disciplines were cardiology (16 CDT, 20%), oncology (10, 13%) and orthopaedics (9, 11%) (appendix p 18).

**Figure 3:**
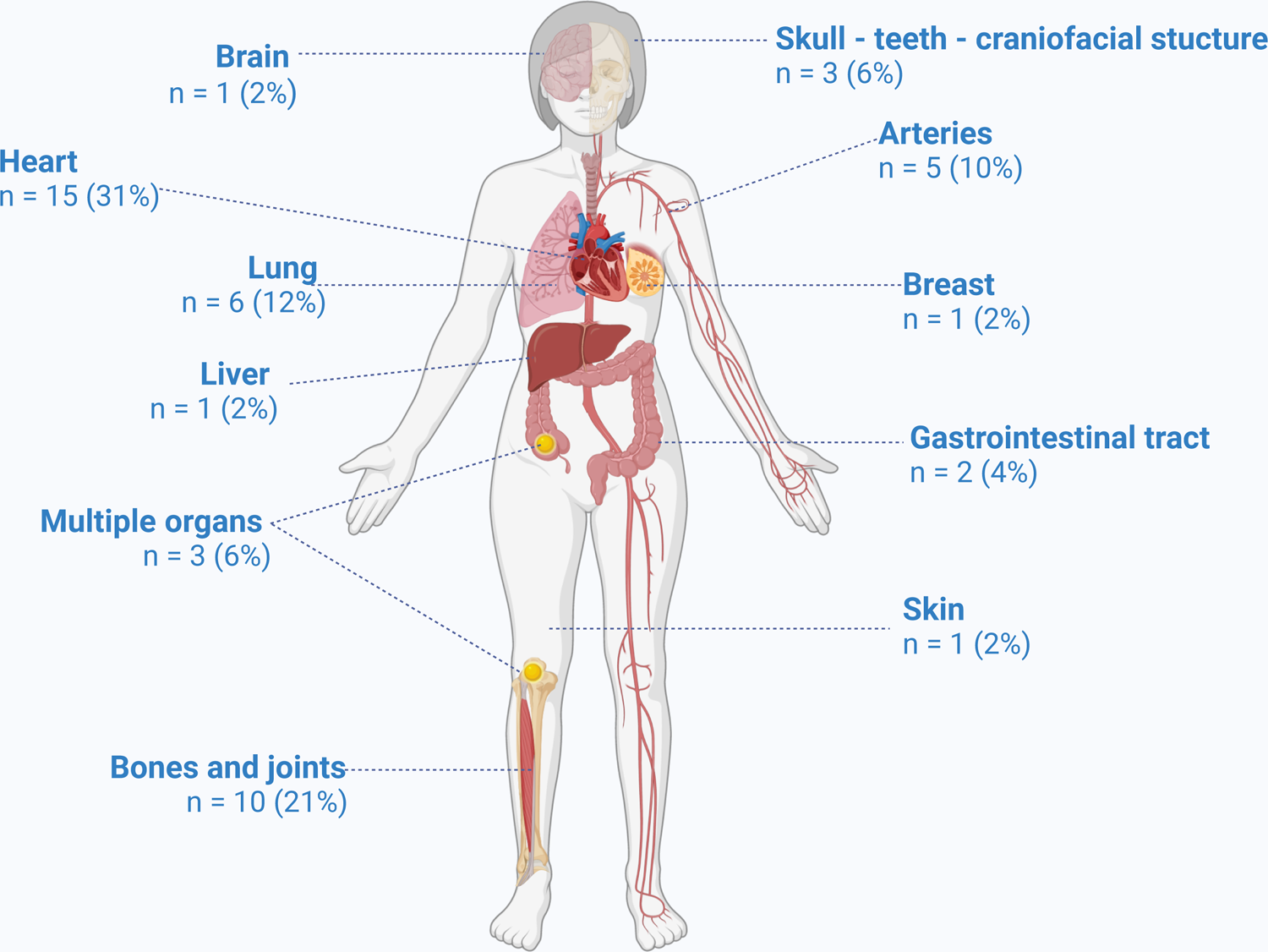
Distribution of organs/systems modelled in the claimed digital twins identified. Created with Biorender. n = 48 claimed digital twin representing specific organs

### Characteristics of the claimed “digital twins”

The patient data used to develop and/or run the 80 unique CDT encompassed: data from clinical notes (15 CDT, 19% of publications), laboratory test results (13 CDT, 16%), medical imaging exams (35 CDT, 44%), wearable device data (12 CDT, 15%), and other data modalities such as ECG, optical tracking, diet data, intra-operative haemodynamic measurements, in 32 CDT (40%) (appendix p 19). Multimodal data incorporation, synthesising various data types and sources, was present in 39 (49%) CDT. Nearly all CDTs (78, 98%) involved data analytics and 29 (36%) some form of advanced visualisation. Eight (10%) CDT involved multiphysics, and nine (11%) were multi-scale. In terms of implementation, 40 (50%) CDT followed a mechanistic approach, 22 (28%) a data-driven approach, 17 (21%) combined both approaches, and one (1%) none of these approaches. Most of the CDT developed (56, 70%) corresponded to models for simulation, and not simulations themselves.

In terms of medical approach, 10 (12%) CDT were categorised as anatomical, 22 (28%) as physiological, and 28 (35%) combined anatomical and physiological features. The objectives of CDTs were prediction (61 CDTs, 76%), simulation (52 CDTs, 65%), monitoring (11 CDTs, 14%), visualisation (10 CDTs, 12%) and generation of synthetic patient data (8 CDTs, 10%). Among the 64 (80% of total) patient-specific CDTs, 44 (69%) were static models, whereas 20 (31%) were designed as dynamic models with regular data inputs or outputs on a daily, hourly, or real-time basis (appendix p 20).

Data flow topology characterises the exchange of information between patient and digital twin. Thirteen (16%) CDTs involved no flow from patient to their virtual counterpart; 58 (73%) used one-way flow from patient to their virtual counterpart; and 9 (11%) enabled two-way data flow between patient and their virtual counterpart (Figure 2). Within the nine two-way digital twins, automated feedback occurred via recommendations to the patient or the physician in six cases and via direct feedback during surgical navigation in three cases.

Finally, regarding the clinical research phase of the CDTs, 78 (98%) were categorised as phase 0 or 1 (pre-clinical phases) and only 2 (2%) involved clinical assessments and thus reached phase 2. No CDT corresponded to phases 3 or 4.

### Unsupervised classification of claimed digital twins

Three clusters were identified using the PAM method (Figure 4, appendix p 21).

**Figure 4:**
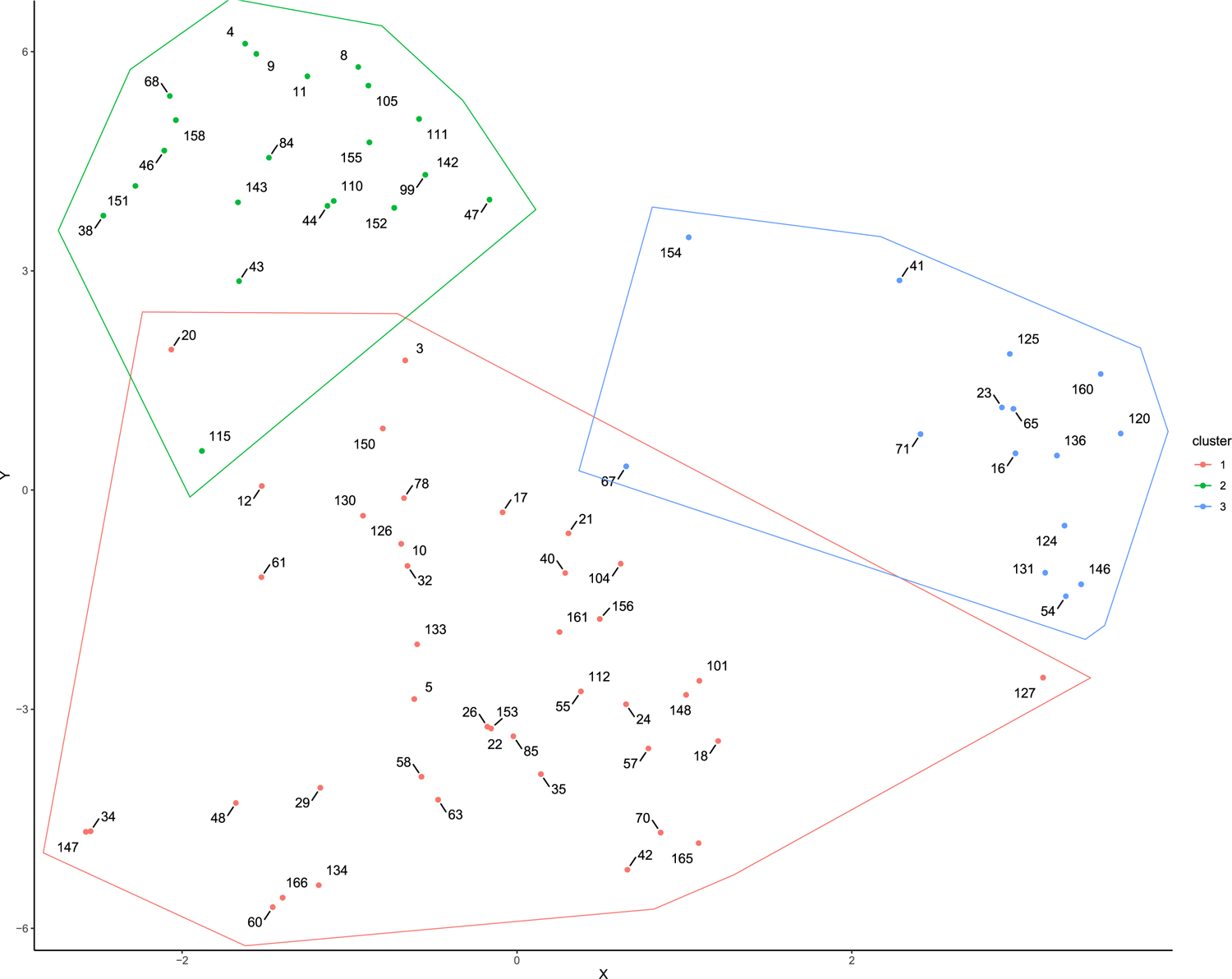
Clusters of claimed digital twins identified. Each point and its associated number correspond to one study and its study identification number.

Cluster 1 included 43 unique CDTs, which corresponded to patient-specific organs or systems models for simulations and predictions. Indeed, 42 (98%) were models to perform simulations, of an organ or system in 42 (98%) cases, and these were anatomical and/or physiological models in 43 (100%) cases relying on a mechanistic approach in 41 (95%) cases. The output of these CDT was patient-specific in 42 (98%) cases, and their use was planned for one-time use in 36 (84%) cases rather than for dynamic use.

Cluster 2 included 22 CDTs and mainly corresponded to prediction models and monitoring systems. In this category, the CDT most often did not involve an anatomical or physiological model of the patient (18 CDTs, 82%). The approach was mainly data-driven (20 CDTs, 91%). These CDTs were often dynamic (13 CDTs, 59%), and provided feedback in 5 (23%) cases.

Cluster 3 included 15 CDTs and usually corresponded to models not linked to patients but designed for research. Indeed, in this group, only one (7%) CDT was patient-specific^63^. Most CDTs were models for simulation (12, 80%) and involved a mechanistic approach (9 CDTs, 60%). Examples of such CDTs are models of the immune response to vaccines,^62^ but also CDTs that corresponded to the generation of synthetic patients for in-silico trials^103^ (5 CDTs, 33%).

## DISCUSSION

This systematic review highlights the growing interest in the concept of patient digital twins while at the same time revealing the lack of a uniform vision of this same concept among research teams.

The publication of scientific articles on digital twins across all sectors has experienced exponential growth, starting from one article in 2014 to more than 1,000 articles per year in 2022, predominantly in the manufacturing industry.^107,108^ Studies focusing on the development and/or evaluation of patient digital twins emerged later, from 2019 onwards, but have seen the same growth, particularly in Europe and North America.

Despite growing interest, there is no consensus definition of the digital twin concept. Depending on the research teams and fields of application, the definitions and characteristics of digital twins differ,^108^ and this is also reflected in our study, with a diversity of definitions and types of CDTs.

If one were to strictly import the digital twin definition used by Michael Grieve^4^ and NASA^5^ from the manufacturing industry and aerospace sector to patients, a digital twin of a patient would be an integrated multi-scale, probabilistic simulation of a patient that uses the best available models, sensor updates, medical history, etc., to mirror the health status of the patient in real-time and act on them. But this definition is not currently achievable since no patient digital twin is able to capture the complexity of the human body in real time. Indeed, creating a patient’s digital twin is different from creating a digital twin of an object like an airplane. An airplane is designed and developed entirely by humans, who have mastery over the composition and physical properties of each part of an airplane, the assembly of all these parts, and the interaction of the whole. In contrast, we still have an imperfect understanding of the functioning of the human body and the interaction of its different organs and systems. It is therefore simpler to model an airplane than the human body with a high degree of fidelity and in a multiscale way. It is also easier to integrate additional sensors and actuators on an airplane’s controls than to have patients continuously wear or have implanted sensors or actuators to obtain the two-way, real-time data exchange between the plane or patient and their digital twin. Moreover, while the digital twin of the airplane relies essentially on the physical properties of the aircraft, the digital twin of a patient will have to go beyond anatomical and physiological models of the patient’s various organs and systems to include models of the patient’s cognitive and emotional functioning.^109^ Finally, concerning the practical use of the terms “patient digital twin”, they must be able to echo patients’ own perceptions of what a digital twin is. These terms are not neologisms, but each refers to concepts already used in everyday language by patients. Because twin refers to a multiple birth, it is likely that the patient pictures a digital twin as a realistic avatar of themselves in the same health state. This implies that in the mind of a clinician or patient not well-versed in computer science, the first representation of a patient digital twin will be that of a 3D representation of the patient’s body and its functioning.

For all these reasons, the concept of a digital twin, which is appropriate in manufacturing, could be considered inappropriate in medicine. However, on the basis of the various articles in this review, we believe that this concept actually corresponds to current advances brought by the multiplication of data sources, data analysis and the visualisation of patients’ state of health. It could also be a useful educational tool for the communication with patients when discussing models and predictions made from their data. Since it is currently not feasible to bring together all the characteristics of the digital twin in manufacturing, we propose the following definition of a patient digital twin: “A patient digital twin is a viewable digital replica of a patient, organ, or biological system that contains multidimensional, patient-specific information”.

The first advantage of this definition is that it can exclude what is not a patient digital twin: generic models of cells, tissues, organs, or biological systems not linked to a patient but used to study disease progression or drug development^29,54,80^; pure cyber-physical systems, that is, systems such as implantable cardioverter-defibrillators, which do not use a representation of the patient and therefore not a “viewable” digital replica of the patient; digital patient data created from patient databases for in silico trials;^47,99,103^ often using generative adversarial networks, which we propose to call “synthetic patients” instead; data sets from another patient, similar to those of the index patient;^77^ machine learning based classifiers, trained on a population to predict a diagnosis;^69^ and patient models built from a single data source, such as demographic characteristics or imaging.^31,47,49^

The second advantage of this definition is that it encompasses the two major trends in patient digital twins revealed by this study. On one hand, digital twins offering a high degree of fidelity, combining advanced anatomical and physiological models, based on mechanistic approaches or combining mechanistic and data-driven approaches.^43,66,105^ However, their limitation is their common reliance on hospital-acquired data such as imaging data (CT, MRI) or intraoperative haemodynamic data. They are therefore generally restricted to one-time purposes, and not in a dynamic form. On the other hand, digital twins corresponding to real-time representations of patients through home-based data collection via wearables, using machine learning techniques for analysis and alert detection.^25,27^ This type of digital twin is dynamic, can be two-way by giving recommendations to patients or by modifying a biological parameter of the patient via medical devices, but often offers little or no information on the patient’s anatomy or physiology. For it to be differentiable from a cyber-physical system or a telemonitoring system, it must integrate different data sources, data analysis systems, and a visualisation of the patient organ, system or body. We thus propose two major categories of patient digital twins (Figure 5):

– Simulation patient digital twins: personalised, viewable digital replicas of patients’ anatomy and physiology based on computational modelling to run simulations predicting outcomes in hypothetical scenarios or evaluating therapeutic approaches. These digital twins are mostly used for one-time assessments rather than continuous monitoring.
– Monitoring patient digital twins: personalised, viewable digital replicas of patients leveraging aggregated health data and analytics to enable continuous predictions of risks and outcomes over time and provide feedback for optimising care. These digital twins are mostly focused on continuous tracking rather than detailed mechanistic simulation.

**Figure 5:**
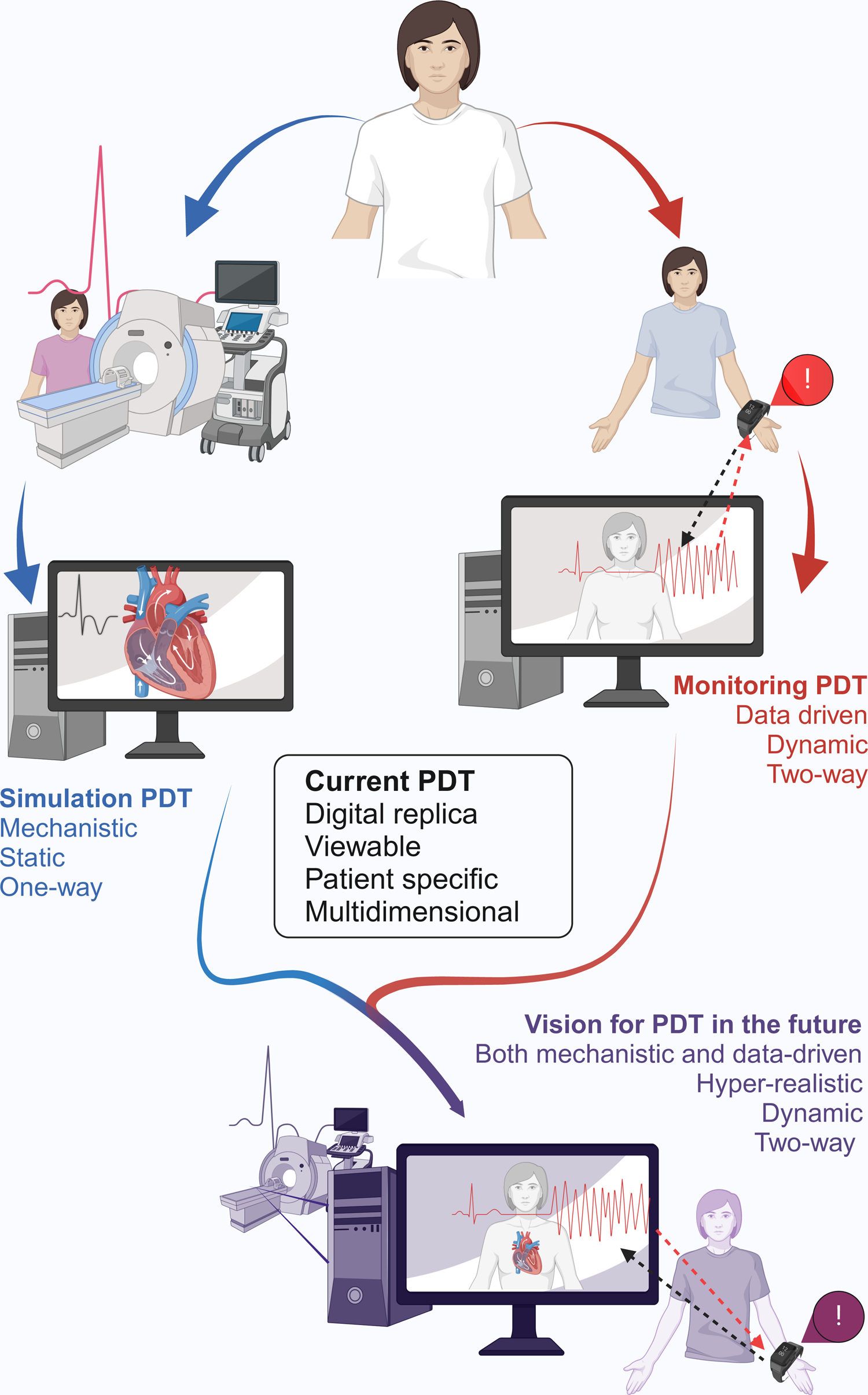
Proposed definitions and characteristics of patient digital twins. Created with Biorender. PDT: Patient digital twin

As technical progress is made, it is likely that these two types of patient digital twins will converge, combining both mechanistic models and increasingly efficient continuous data collection and data-driven approaches.^110^ However, currently, only 14 CDTs from this review would meet the definition of patient digital twins combining multidimensional, patient-specific data, data analytics and advanced visualisation, all being simulation patient digital twins.^23,34,37,43,48,57,58,61,66,70,88,98,105,106^ Digital twins for surgical navigation systems are the most advanced, with three teams presenting patient digital twins combining both a mechanistic approach and dynamic real-time adaptation.^58,88,106^

The main benefit expected from these patient digital twins is to offer increasingly personalised medicine, taking into account all the available data and patient-specific simulations and predictions. This would make it possible, for example, to choose the most appropriate drug treatment based on the patient’s medical history, allergies, comorbidities and genetic profile;^22^ the most appropriate stent for the configuration of the aortic dilatation;^23^ and the least risky surgical approach thanks to pre-operative simulations. But the benefits could also be for patient education and engagement: patients would interact with their digital replica to better understand their own body, health conditions, and influence of behaviours, and collaborate more with providers.

### Strengths and limitations

The strength of this review is to be the first to have carried out a systematic analysis of the literature concerning patient digital twins, and to have been able to identify the major trends. We also acknowledge several limitations. The main one is that in the absence of a consensual definition of the digital twin concept, we chose to include all articles in which the authors claimed they had developed a digital twin or part of a digital twin. This has undoubtedly led us to not include some articles that meet the concept of a digital twin but did not use that term, and to include some articles that in fact did not develop a digital twin but only a simple prediction model. Another limitation is that we realised that some characteristics of digital twins were not appropriate either because very industry-specific such as the “multiphysic” characteristic or because they did not allow us to be consistent in our assessment. For example, we often failed to determine whether the presented digital twin was at phase 0 or 1 of development and chose to merge these two categories as preclinical. Finally, we did not include articles written in languages other than English, French, or German, and this may have led to underrepresenting the work of some regions of the globe.

## CONCLUSIONS

In conclusion, we propose that a patient digital twin be defined as “a viewable digital replica of a patient, organ, or biological system that contains multidimensional, patient-specific information”. We currently identify two categories, simulation patient digital twins and monitoring patient digital twins. In the future, we envisage a fusion of these two types of digital twins that will combine a high degree of fidelity based on anatomical and physiological models with real-time updating and feedback to the patient.

## Contributors

DD led all aspects of the Review: conceptualisation, data curation (title and abstract screening, full-text screening, and data extraction), formal analysis, project administration, supervision, validation, and writing (original draft, review, and editing); AG was involved in conceptualisation, data curation (title and abstract screening, full-text screening, and data extraction), formal analysis, validation, and writing (review, and editing). All authors have had full access to the data and accept responsibility to submit for publication.

## Data Availability

All data produced in the present work are contained in the manuscript

## Acknowledgments

The authors wish to acknowledge the use of Claude 2.1, an artificial intelligence assistant created by Anthropic, for the translation of selected passages from French to English and the rewriting of these passages into academic English.

