## Supplementary appendix for "Patient digital twins: an introduction based on a scoping review"

**Supplementary material table of contents**

**Full search strategies**

**EMBASE via OVID**

1. exp digital twin/
2. ("digital" and "twin*").ab,ti.
3. exp medicine/
4. medic*.ab,ti.
5. exp surgery/
6. surg*.ab,ti.
7. exp radiology/
8. exp radiography/
9. radiolog*.ab,ti.
10. exp diseases/
11. diseas*.ab,ti.
12. exp diagnosis/
13. diagnos*.ab,ti.
14. exp therapy/
15. treat*.ab,ti.
16. exp prognosis/
17. prognos*.ab,ti.
18. exp health/
19. health*.ab,ti.
20. exp patient/
21. patient*.ab,ti.
22. exp personalized medicine/
23. "precision medicine".ab,ti.
24. "personalized medicine".ab,ti.
25. "personalised medicine".ab,ti.
26. 1 or 2
27. 3 or 4 or 5 or 6 or 7 or 8 or 9 or 10 or 11 or 12 or 13 or 14 or 15 or 16 or 17 or 18 or 19 or 20 or 21 or 22 or 23 or 24 or 25
28. 26 and 27

**Google scholar**

digital twin medicine OR surgery OR radiology OR disease OR diagnosis OR treatment OR prognosis OR health OR patient OR "precision medicine" OR "personalised medicine" OR "personalized medicine"

**IEEE**

(Abstract:"digital" AND Abstract:"twin*") AND ("medic*" OR "surg*" OR "radiol*" OR "disease*" OR "treat*" OR "prognos*" OR "health*")

*AND (research made in two times because IEEE allow only a finite number of search terms)*

(Abstract:"digital" AND Abstract:"twin*") AND ("patient*" OR "precision medicine" OR "personalised medicine" OR "personalized medicine")

**MEDLINE via Pubmed**

"digital"[Title/Abstract] AND "twin*"[Title/Abstract] AND ("medicine"[MeSH Terms] OR "medic*"[All Fields] OR ("surgery"[MeSH Subheading] OR "surg*"[All Fields]) OR ("radiology"[MeSH Terms] OR "radiography"[MeSH Terms] OR "radiolog*"[All Fields]) OR ("disease"[MeSH Terms] OR "diseas*"[All Fields]) OR ("diagnosis"[MeSH Terms] OR "diagnos*"[All Fields]) OR ("therapeutics"[MeSH Terms] OR "treat*"[All Fields]) OR ("prognosis"[MeSH Terms] OR "prognos*"[All Fields]) OR ("health"[MeSH Terms] OR "health*"[All Fields]) OR ("patients"[MeSH Terms] OR ("patient s"[All Fields] OR "patients"[MeSH Terms] OR "patients"[All Fields] OR "patient"[All Fields] OR "patients s"[All Fields])) OR ("precision medicine"[MeSH Terms] OR ("precision medicine"[MeSH Terms] OR ("precision"[All Fields] AND "medicine"[All Fields]) OR "precision medicine"[All Fields] OR ("personalised"[All Fields] AND "medicine"[All Fields]) OR "personalised medicine"[All Fields]) OR ("precision medicine"[MeSH Terms] OR ("precision"[All Fields] AND "medicine"[All Fields]) OR "precision medicine"[All Fields] OR ("personalized"[All Fields] AND "medicine"[All Fields]) OR "personalized medicine"[All Fields])))

**Scopus**
TITLE-ABS-KEY ( "digital" AND "twin*" )

AND

( TITLE-ABS-KEY ( medic* ) OR TITLE-ABS-KEY ( surg* ) OR TITLE-ABS-KEY ( radiolog* ) OR TITLE-ABS-KEY ( disease* ) OR TITLE-ABS-KEY ( diagnos* ) OR TITLE-ABS-KEY ( treat* ) OR TITLE-ABS-KEY ( prognos* ) OR TITLE-ABS-KEY ( health* ) OR TITLE-ABS-KEY ( patient* ) OR TITLE-ABS-KEY ( "precision medicine" ) OR TITLE-ABS-KEY ( "personalised medicine" ) OR TITLE-ABS-KEY ( "personalized medicine" ) )

**Web of science:**
1: TS=("digital" and "twin*") OR TI=("digital" and "twin*")
2: (TS=("medic*")) OR TI=("medic*")
3: (TS=("surg*")) OR TI=("surg*")
4: (TS=("radiolog*")) OR TI=("radiolog*")
5: (TS=("disease*")) OR TI=("disease*")
6: (TS=("diagnos*")) OR TI=("diagnos*")
7: (TS=("treat*")) OR TI=("treat*")
8: (TS=("prognos*")) OR TI=("prognos*")
9: (TS=("health*")) OR TI=("health*")
10: (TS=("patient*")) OR TI=("patient*")
11: (TS=("precision medicine")) OR TI=("precision medicine")
12: (TS=("personalised medicine")) OR TI=("personalised medicine")
13: (TS=("personalized medicine")) OR TI=("personalized medicine")
14: #13 OR #12 OR #11 OR #10 OR #9 OR #8 OR #7 OR #6 OR #5 OR #4 OR #3 OR #2
15: #14 AND #1

**Final data charting**

| Variable | Explanation |
|---|---|
| Study ID | Study ID attributed by our team |
| Reference | PMID reference or DOI if PMID not available |
| Author | First author name |
| Year | Year of publication |
| Country | Country of the corresponding author |
| Unique | Yes: unique digital twin; no: the same digital twin was already used in a previous study included in this review |
| Definition | Definition of digital twin by the study authors |
| Definition: yes or no | Yes: the authors provided a definition of digital twin |
| Definition: digital replica | Yes: the authors refer to a digital replica, virtual counterpart, or similar description in their definition of digital twin |
| Definition: high fidelity | Yes: the authors refer to an accurate, high-fidelity or similar description in their definition of digital twin |
| Definition: multiscale | Yes: the authors refer to a multiscale or combination of information at different levels (e.g. cellular, tissue, and organ) in their definition of digital twin. |
| Definition: two ways | Yes: the authors refer to a two-way communication between the physical and the digital twin, or include the notion of feedback to the physical object in their definition of digital twin |
| Definition: real time | Yes: the authors refer to real-time update, or continuous stream of information from the physical to the digital twin in their definition of digital twin. |
| Definition: patient specific | Yes: the authors refer to a patient in their definition of digital twin. |
| Type: System/organ* | Organ: digital twin of an organ or part of the body (e.g. vertebra)<br>System: digital twin of a biological system (e.g. immune system)<br>Other: prediction models, pharmacological models, etc. |
| Medical discipline | Medical discipline of the digital twin (e.g. cardiology for digital twins of the heart) |
| Data: notes | Yes: the digital twin presented required patient data from clinical notes (demographics or medical history) to be developed and/or used |
| Data: laboratory | Yes: the digital twin presented required patient data from laboratory results (e.g. results from blood tests) to be developed and/or used |
| Data: imaging | Yes: the digital twin presented required patient data from imaging (e.g. CT, MRI images) to be developed and/or used |
| Data: wearable | Yes: the digital twin presented required patient data from wearable devices (e.g. activity trackers, continuous glucose monitoring) to be developed and/or used |
| Data: other | Yes: the digital twin presented required patient data from other sources (e.g. ECG data, optical tracking data, diet data) to be developed and/or used. |
| Multimodal data* | Yes: different types and/or sources of patient data were required to develop and/or use the digital twin presented (e.g. clinical and laboratory data, or CT and MRI imaging data). |
| Model type* | Anatomical: the digital twin presented involves only anatomical data, usually a 3D representation of the organ.<br>Physiological: the digital twin presented involves only physiological processes.<br>Both: the digital twin presented combines anatomical and physiological models<br>Other: the digital twin presented do not involve an anatomical or physiological model. Often it refers to a prediction model using data-driven approaches. |
| Multiphysics* | Yes: at least two physics disciplines were involved in the development of the digital twin presented. |
| Multiscale* | Yes: at least two scales are involved in the digital twin presented among molecular, cellular, tissue, organ, system, patient |
| Approach* | Mechanistic: An approach that uses mathematical equations and scientific first principles to explicitly model expected relationships between system variables.<br>Data-driven: An approach that applies machine learning algorithms to derive predictive models purely from patterns in collected data.<br>Both: An approach that integrates mechanistic frameworks with data-driven optimization of model parameters<br>Other: No such approaches were used (e.g. visualisation only) |
| Model for simulation* | Yes: The digital twin presented represents an entity (organ, biological system) that can be used to simulate different states.<br>No: Otherwise. For example - monitoring systems relying on wearable devices and alerts. |
| Intent to update DT* | The authors intention is to develop a digital twin which is:<br>Static: for a one-time use with the patient data<br>Dynamic: for regular use with the patient data<br>Not patient specific: no update is possible because no patient data are linked. |
| Frequency of data update | For dynamic digital twins, the frequency of update of data entry: in real-time (several times per second), hour (several times per hours), day (several times per day), not specified. |

| | |
|---|---|
| Frequency of output update | For dynamic digital twins, the frequency of the digital twin output (e.g. recommendation, haptic feedback during surgery): in real-time (several times per second), hour (several times per hours), day (several times per day), not specified. |
| DT: one way/two way* | No: no communication between the physical and digital twin<br>One: communication from the physical to the digital twin, but not the reverse.<br>Two: two-way communication between the physical and the digital twin. |
| DT: feedback | Type of feedback provided by two-ways digital twin systems<br>Recommendation: for physician or patient<br>Surgical navigation: haptic feedback, visual feedback, etc. |
| DT: patient specific output* | Yes: the digital twin presented is patient-specific, i.e. its output is specific to a patient or intended to be specific to a patient. |
| DT: phase | Clinical research phase based on[1] :<br>0 (discovery/invention): prototype design<br>1 (safety/dosage): in-silico optimization and usability testing<br>2 (efficacy/side-effects): clinical assessments<br>3 (efficacy): clinical trials<br>4 (safety/effectiveness): post-deployment surveillance |
| Advanced analytics* | Yes: the digital twin presented involved data analytics |
| Advanced visualisation* | Yes: the digital twin presented involved some form of advanced visualisation, i.e. the digital replica of the cell, tissue, organ, system or patient can be “viewable” in 2D or 3D |
| Objective: simulation* | Yes: one of the objectives of the digital twin presented is simulation |
| Objective: prediction* | Yes: one of the objectives of the digital twin presented is prediction |
| Objective: monitoring* | Yes: one of the objectives of the digital twin presented is monitoring |
| Objective: visualisation* | Yes: one of the objectives of the digital twin presented is visualisation |
| Objective: generation* | Yes: one of the objectives of the digital twin presented is to generate synthetic patient data |

* Variables that were included for the clustering of the different digital twins.

1 Park Y, Jackson GP, Foreman MA, Gruen D, Hu J, Das AK. Evaluating artificial intelligence in medicine: phases of clinical research. *JAMIA Open* 2020; **3**: 326–31.

**List of studies excluded at the full-stage screening stage and reasons.**

**List of records not retrievable.**

1. Díaz RG, Laamarti F, El Saddik A. DTCoach: Your Digital Twin Coach on the Edge During COVID-19 and Beyond. IEEE Instrum Meas Mag. 2021 Sep;24(6):22–8.

2. Li X. Real-time digital twins end-to-end multi-branch object detection with feature level selection for healthcare. J Real-Time Image Process. 2022 Oct 1;19(5):921–30.

3. Wang W, He Y, Li F, Li J, Liu J, Wu X. Digital twin rehabilitation system based on self-balancing lower limb exoskeleton. Technol Health Care Off J Eur Soc Eng Med. 2023;31(1):103–15.

4. Marty P, Boehm C, Paverd C, Rominger M, Fichtner A. Full-waveform ultrasound modeling of soft tissue-bone interactions using conforming hexahedral meshes. In: Medical Imaging 2022: Physics of Medical Imaging [Internet]. SPIE; 2022 [cited 2024 Jan 25]. p. 877–91. Available from: https://www.spiedigitallibrary.org/conference-proceedings-of-spie/12031/120313H/Full-waveform-ultrasound-modeling-of-soft-tissue-bone-interactions-using/10.1117/12.2611548.full

**List of excluded articles at full-text screening stage.**

Wrong study type (conference paper, commentary, book chapter, abstract)

Retracted articles

22. Jiang J, Li Q, Yang F. TCM Physical Health Management Training and Nursing Effect Evaluation Based on Digital Twin. Sci Program. 2022 Sep 27;2022:e3907481.

23. Chen J. 3D Visualization Analysis of Motion Trajectory of Knee Joint in Sports Training Based on Digital Twin. Comput Intell Neurosci. 2022;2022:3988166.

Articles not peer-reviewed

24. Cockrell C, Schobel-McHugh S, Lisboa F, Vodovotz Y, An G. Generating synthetic data with a mechanism-based Critical Illness Digital Twin: Demonstration for Post Traumatic Acute Respiratory Distress Syndrome [Internet]. bioRxiv; 2022 [cited 2024 Jan 25]. p. 2022.11.22.517524. Available from: https://www.biorxiv.org/content/10.1101/2022.11.22.517524v1

25. Zhang Y, Zhang K, Prakosa A, James C, Zimmerman SL, Carrick R, et al. Predicting Ventricular Tachycardia Circuits in Patients with Arrhythmogenic Right Ventricular Cardiomyopathy using Genotype-specific Heart Digital Twins. MedRxiv Prepr Serv Health Sci. 2023 Aug 1;2023.05.31.23290587.

26. Herrgårdh T, Simonsson C, Ekstedt M, Lundberg P, Stenkula KG, Nyman E, et al. A multi-scale digital twin for adiposity-driven insulin resistance in humans: diet and drug effects. Diabetol Metab Syndr. 2023 Dec 4;15(1):250.

No claimed digital twin

Not a patient digital twin

Development limited to a framework or a technology

**Number of studies claiming to be developing or to have developed/evaluated a patient digital twin published per year.**

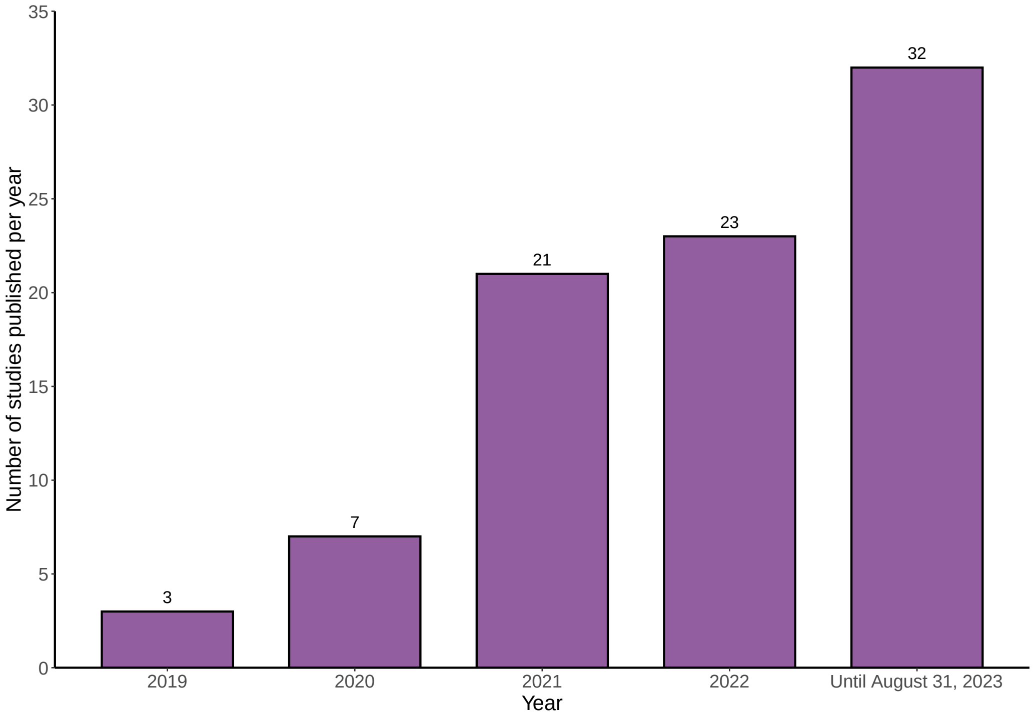

**Geographic distribution of studies claiming to be developing or to have developed/evaluated a patient digital twin.**

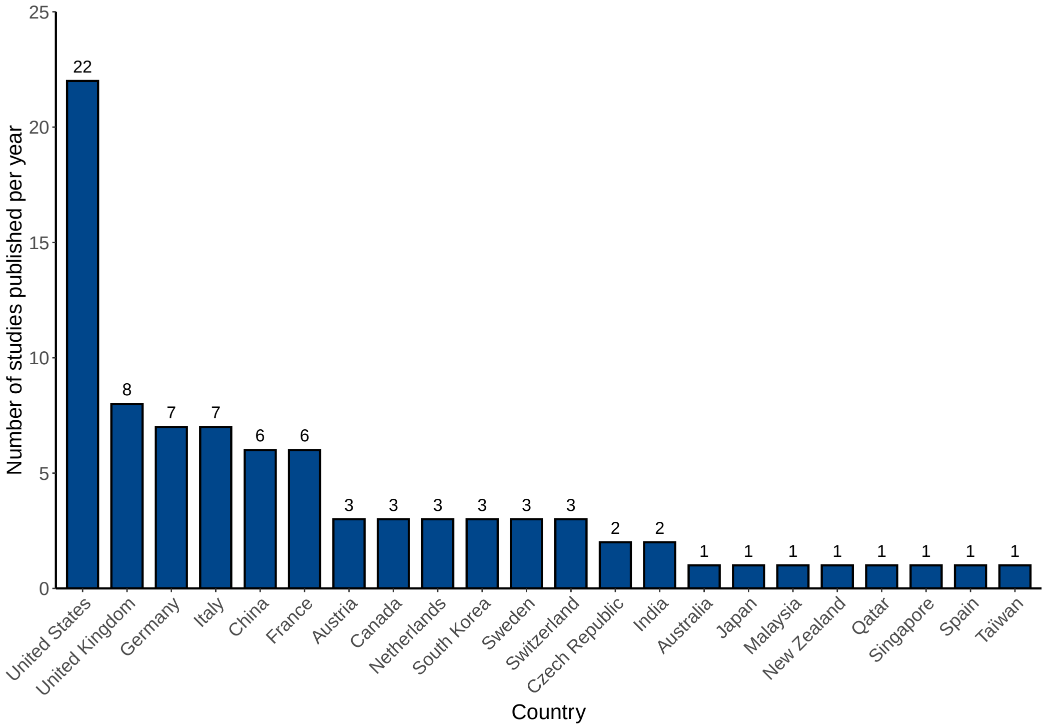

**Definitions of digital twin provided by authors.**

| Study ID | Reference | Author | Year | Definition of digital twin provided by authors |
|---|---|---|---|---|
| 3 | 30648344 | Chaksu | 2019 | The digital twin concept can be broadly described by three characterisations: active, where a digital replica (digital twin) of a physical system (physical twin) is continuously updated by information and data collected from the physical twin; semi-active, where time-varying data are collected, but rather than performing a continuous update, the information is analysed after the data are collected; and passive, where the digital twin utilises measurements from a physical twin which are not continuously updated, which may include some modelling assumptions |
| 4 | 10.1109/ACCESS.2019.2909828 | Liu | 2019 | At present, one commonly used defnition from NASA is, "A Digital Twin is an integrated Multiphysics, multiscale, probabilistic simulation of an as-built vehicle or system that uses the best available physical models, sensor updates, fleet history, etc., to mirror the life of its corresponding flying twin.'" A digital twin consists of three parts: physical objects, virtual objects, and connected data. From the description of the digital twin, it has the following characteristics :(1) Real-time reflection: Various types of physical object data should be integrated, and it can keep real-time mapping of physical objects. (2) Interaction and convergence: It exists in and co-evolves with the full lifecycle of physical objects. (3) Evolution and iteration: It can not only describe physical objects, but also optimize physical objects based on the iterative virtual model. DTH is a new medical simulation approach for a medical activity or a medical system to provide fast, accurate and efficient medical services using DT technology with multiscience, multi-physics and multi-scale models. DTH mainly consists of three parts: physical object, virtual object and healthcare data. The physical object may be a medical device or a wearable device for the elderly, a patient, an external factor such as social behavior, weather or government policy influencing their health, or a system consisting of several or all of these unit objects. The virtual object is the medical device model, wearable device model, digital person model, external factor model, and the digital system model, respectively. The healthcare data includes detection data from medical devices or external systems, real-time monitoring data from wearable devices, simulation data from digital models, historical data and medical records from medical institutions, and service data from service platforms or systems that connect the physical and virtual spaces. The operation mechanism of DTH consists of four main stages. First, accurate digital twin models should be established corresponding to the physical entity objects (unit level object or system level object) by means of advanced modeling techniques or tools such as SysML, Modelica, SolidWorks, 3DMAX and AutoCAD. Second, data connection should be done through health IoT and mobile internet technologies in order to maintain the interaction between physical and virtual objects in real time. Third, simulation is validated by quick execution and calibration to ensure that the model is correct. Fourth, according to the needs and actual conditions, model evolution should be carried out continuously to optimize and iterate the DT models. Then, data from DT models are fedback to the physical objects and the service systems to provide better solutions for elderly healthcare services. |
| 5 | 10.1007/s00772-019-00569-4 | Hemmler | 2019 | The digital twin, as a digital replica of this real entity |
| 8 | 31267697 | Hirschvogel | 2020 | DT is defined as the virtual (digital) counterpart of a physical system. The DT starts living as the physical is being prototyped, dies when it is disassembled, and follows its physical twin during its whole life, resembling its status and being continuously connected and synchronized with it. Thanks to AI, the DT continuously controls and monitors its physical twin status, with the aim of optimizing its performance by triggering self-optimization and self-healing mechanisms. The digital-physical twins interaction is based on a ``closed-loop", which refers to the continuous exchange of data between the cyber and physical worlds in order to continuously optimize the physical side. receives data from its physical twin, reconfigures itself to be synchronized with it, applies AI algorithms to detect anomalies and then suggests self-healing or optimization actions. Thanks to the DT, all stakeholders have a quick and easy access to the physical object status, to monitor and control it and eventually trigger required actions. The success of DT technology in manufacturing, documented in the extensive survey reported in [4], has motivated several studies aimed at extending it to humans, by designing, human DTs, that is computer models of humans tailored to any patient to allow researchers and clinicians to monitor the patient's health, for providing and test treatment protocols. Human DTs differ from DTs developed and used in Industry 4.0 because they are not continuously connected to their physical twin. In this case, users and experts are meant to continuously update the DT with the input of medical digital, data describing the health condition of the physical twin. |
| 9 | 10.1109/ACCESS.2020.2971576 | Barricelli | 2020 | The digital twin concept was proposed in 2002 and referred to the digital replica of a physical object. A new vision of the digital twin was introduced in to re-de ne it as a digital replication of a living or nonliving physical entity. By bridging the |

| | | | | |
|---|---|---|---|---|
| | | | | physical and virtual worlds, data are transmitted seamlessly, which enables the virtual entity to exist simultaneously with the physical entity. Digital twin facilitates the means to monitor, understand, and optimize the functions of the physical entity and provides continuous feedback to improve the quality of life and well-being. A digital twin is hence the convergence of several technologies, such as data analytics and artificial intelligence, haptics and the Internet of Things, data visualization techniques, cybersecurity and communication networks. The digital twin, as defined in [2], has multiple characteristics. Other than a unique identifier, the digital twin uses sensors and actuators, which enables it to continuously collect data and renders it an accurate replica of the real twin at any given time, as well as conveys feedback to the real twin. These sensors and actuators can have the form of wearables and personal health devices, which use has exploded in recent years. In this paper, we use the digital twin in its more recent and much broader definition, which considers the digital twin as a replica of any living or nonliving entity. We use sensors for the digital twin as humans use their five senses. These sensors are used by the digital twin to learn as much information as available about the real twin and his or her context. These collected data are then stored in cloud storage, which serves as the ``memory" of the digital twin. Data processing by means of artificial intelligence algorithms and data analytics is the mapping for the digital twin of human intelligence. This process enables decision-making and feedback transmission to the real twin. Feedback can take the form of actuation, such as haptic actuation or notifications or manipulation of a home environment to improve the health and well-being of the real twin. |
| 10 | 33225302 | Lal | 2020 | Digital twin allows clinical or research interventions to be tested first in a virtual environment, minimizing safety concerns to real patients |
| 12 | 10.1016/j.bspc.2019.101813 | Goodwin | 2020 | A digital twin associated with a physical system is a mathematical model that represents the system. |
| 16 | 10.1007/s41745-020-00185-2 | Subramian | 2020 | Digital replicas of processes, systems or devices used for understanding study or prediction |
| 17 | 10.3233/FI-2020-1943 | Stefano | 2020 | A digital representation of that patient physiology. |
| 18 | 33651703 | Fu | 2021 | Digital representations of clinical cases |
| 20 | 10.1109/ACCESS.2021.3063786 | Béthencout | 2021 | A mathematical model of the ``(biological stimulus)-(sensor)-(processor)" system, whose computer implementation defines a digital twin that can generate, based on physical input parameters, a wide variety of physically plausible situations. |
| 21 | 33360683 | Zhou | 2021 | Patient-specific digital twins or virtual patients built on clinical data |
| 22 | 34271532 | Camps | 2021 | A comprehensive virtual tool that coherently integrates a patient's clinical data with mechanistic physiological knowledge and that can inform therapeutic and diagnostic decision-making through simulations. |
| 23 | 34603366 | Barbiero | 2021 | A virtual prototype of patients mirroring the underlying biological system (Gelernter, 1993; Laubenbacher et al., 2021) combining information at organ, tissue, and cellular level. |
| 29 | 34351462 | Hernigou | 2021 | Near real-time digital image of a physical human |
| 34 | 33805617 | Cho | 2021 | Data reconstructed in the virtual environment to reproduce reality, which provides answers to a range of clinical questions |
| 35 | 33975097 | Gilette | 2021 | Digital replicas of patient [hearts] derived from clinical data that match like-for-like all available clinical observations. |
| 37 | 34431016 | Gilette | 2021 | Digital replicas of a patient that match all available clinical observations with high fidelity |
| 38 | 10.1186/s13634-020-00715-1 | Conte Alcaraz | 2021 | Digital counterpart of body |
| 41 | 10.3390/electronics10182220 | Gonzalze-Abril | 2021 | Dynamic digital representation of the patient's anatomy and physiology through computational models which are continuously updated from clinical data. (...) Highly capable simulation models, especially those that consume streaming data from sensors to anticipate maintenance issues, impending failure of components and improving performance. |
| 44 | 10.3390/app11125576 | Allen | 2021 | A digital twin of a patient is a simulation of the patient's trajectory that behaves identically to the patient in terms of outcomes. |
| 46 | 10.1109/JIOT.2021.3051158 | Elayan | 2021 | A virtual replica of a physical asset that reflects the current status through real-time transformed data. (...) The concept of this technology refers to a digital replica of the physical object. DT combines Artificial Intelligence (AI), Data Analyt ics, IoT, Virtual and Augmented Reality paired with digital and physical objects. This integration allows real-time data analysis, status monitoring to head off problems before they even occur, risk management, cost reduction, and future opportunities prediction |
| 47 | 35442211 | Tardini | 2022 | A digital replica (digital twin) of a physical entity or process is virtually recreated, with similar elements and dynamics, to perform real-time optimization and testing. In health care, coupled digital twins, that is, digital twin dyads, could be created for both patients and for the therapy process and used to inform in a quantitative manner adaptive therapy decision- |

| | | | | |
|---|---|---|---|---|
| | | | | making and allow personalization and optimization of health outcomes, prediction and prevention of adverse events, and planning interventions |
| 48 | 10.1109/JIOT.2022.3176300 | Tai | 2022 | A digital twin combines various advanced technologies to create actual mapped devices in the virtual space, reflecting the relevant physical devices throughout the life cycle process. Continuous fine-tuning and optimization are performed to optimize the actual devices. |
| 55 | 35138455 | Hernigou | 2022 | Concept for modeling and integrating physics and heterogeneity, allowing simulation and optimization with multiscale systems. |
| 57 | 35403831 | Ahmadian | 2022 | The in-silico representation of a complex system which serves as a virtual model to analyze the current condition and predict its future performance based on real-time data characterizing the system condition. |
| 60 | 10.3390/machines10070530 | Quin | 2022 | The virtual analogy of the physical object, which can be used to simulate, analyze, and monitor the physical object |
| 61 | 35581423 | Venkatapurapu | 2022 | An instance of the model that mimics a real patient |
| 63 | 10.3390/electronics11030502 | Poletti | 2022 | Virtual representation of a physical product […] developed to monitor real-time the product and obtain virtual insights and data for the optimization of design or control processes. |
| 65 | 10.1016/j.cma.2022.115315 | Zohdi | 2022 | Key tools include digital-twins, which are digital replicas of complex systems which can be safely manipulated and optimized in a virtual world and then deployed in the physical world afterwards, reducing costs of experiments and accelerating development of new technologies. This process involves the application of machine-learning to digital-twins, whereby they learn from their mistakes/errors and constantly evolve to improve in a virtual environment. Digital-twins can also run in tandem with real systems and thereby serve as controllers. |
| 68 | 10.1109/ACCESS.2022.3145984 | Sharotry | 2022 | Virtual and precise depiction of a physical system, allowing real-time analysis for detection, prediction, prevention, and optimization to increase productivity |
| 69 | 10.1080/09540091.2021.2013443 | Zhang and Tai | 2022 | Virtual replicas of many physical entities, such as systems, devices, people and even places. |
| 70 | 35295404 | Jung | 2022 | Models that replicate all available observations with high accuracy |
| 71 | 10.1186/s13073-022-01048-4 | Li | 2022 | An in silico model that brings together the technology to map, monitor, and control real-world entities by continually receiving and integrating data from the physical twin to provide an up-to-date digital representation of the physical entity |
| 78 | 36094958 | Silfvergren | 2022 | Digital twins come in different categories, e.g. online twins, which are updated in real time by connecting with a sensor, and offline twins, which are only occasionally updated using new data. Another distinction is between black box twins, utilising machine learning and statistical models, and physiologically based twins. A physiologically based digital twin is a personalised computer model that describes the underlying physiology in a specific person or patient |
| 84 | 36121054 | Chakshu | 2022 | A human digital-twin is one such model, widely used in other areas,23–25 which is a digital replica of a human system or sub-system. This replica is a personalised digital representation, in terms of structure or functioning or both, of an individual or patient's system. It can provide real-time feedback on how a patient's health is likely to vary based on their current known condition using periodic input data from the patient's vitals |
| 104 | 36191352 | Ang | 2022 | Digital copy of a physical system capable of accurately simulating, replicating, and predicting the behaviour of the physical system in various scenarios |
| 105 | 36560115 | Al-Zyoud | 2022 | The virtual representation of a living entity (e.g., human beings, animals, plants, etc.) or nonliving entity (e.g., business model, product, process, system, event, machine, building, etc.) that allows real-time interaction and communication between both the real twin and the digital twin to help with the modelling, monitoring, understanding, and optimization of the functions and behaviour of the real twin |
| 109 | 36897525 | Bahrami | 2023 | A virtual presentation of the real-world patient that contains all his/hers organs and processes, and it is connected to the real world by sensor data or patient feedback. |
| 110 | 10.1038/s41598-023-43618-5 | Cen | 2023 | Virtual mirror of ourselves that allows us to simulate our personal medical history and state of health using data-driven analytical algorithms and theory-driven physical knowledge |
| 115 | 36983097 | Thamotharan | 2023 | The digital twin (DT) is a framework that could fuse multiple models, data, and provide interfaces to virtualize physical entities with different viewpoints. In essence, the DT creates a digital replica of a physical object, process, or system orchestrated through a suite of models that exchange information with physical entities in real-time. The DT integrates these technologies appropriately for real-time data aggregation, system analysis, status monitoring, knowledge creation, providing deeper insights, risk management, bespoke displays, informed actions, agile situation management strategies, prediction, and others. The health digital twin (HDT) has three parts: patient (PT), virtual digital twin (VT), and PT–VT |

| | | | | |
|---|---|---|---|---|
| | | | | interactions. However, orchestrating an HDT requires novel data sources, new models, and techniques to achieve personalized management. The main advantage of an HDT is its ability to handle both historical and real-time data that is missing in current approaches. With an HDT, management could be tailored to focus on a particular individual's contextual data. |
| 125 | 37488327 | Chasseloup | 2023 | A way to mirror a real life system (e.g., a plane or any part of it such as an engine). The model integrates historical and real time data to enable informed and tailored predictions under multiple conditions. |
| 127 | 10.1109/TIM.2023.3298389 | Zhu | 2023 | A virtual replica of physical objects. Digital twin technology, which uses real-world data to create a virtual replica of physical objects, has been rapidly growing in popularity, particularly in the medical field |
| 142 | 37106642 | Moztarzadeh | 2023 | Digital twin is a virtual replica of a physical object that is connected to the original through a system of operations. The virtual twin should closely mimic the behavior of the physical object, providing real-time data and allowing for the timely identification and resolution of errors. Ultimately, the digital twin should be able to sustain itself and improve the physical model through the incorporation of digital intelligence and iterative improvement. A healthcare digital twin is a virtual representation of a person that utilizes a lifetime of data and AI-powered models to predict the individual's health status and provide recommendations for clinical questions. These digital replicas can be used to predict the outcome of certain procedures, such as lung cancer diagnosis |
| 143 | 37189587 | Moztarzadeh | 2023 | Digital representations of physical objects or systems, providing a way to monitor, control, and optimize their performance in real time |
| 146 | 37217483 | Viola | 2023 | The generation of synthetic data by high-fidelity computer models might be an effective strategy to mitigate the above issues and this is one of the main aims of digital medicine. In fact, these models are referred to as digital twins and, when provided with appropriate input parameters, they can be used to surrogate real patients with 'on demand' features. |
| 147 | 37391453 | Lee | 2023 | Data that is recreated in a virtual setting to mimic reality. It can help develop and select the most efficacious treatment options for each patient and predict the outcomes |
| 150 | 37008895 | Batagov | 2023 | A mechanistic numerical model of a particular patient calibrated to the individual's phenotypic and clinical data at a particular time point. |
| 151 | 37549083 | Sarp | 2023 | A virtual replica |
| 153 | 37392493 | Azzolin | 2023 | Digital replicas of patient hearts systematically integrating clinical data that match like-for-like all available clinical observations |
| 155 | 10.1016/j.knosys.2022.110138 | Manocha | 2023 | Refers to the digital replication of the actual object. DT has the capability to combine physical objects and their digital replica with different modern techniques such as IoT, Artificial Intelligence (AI), virtual reality, and many others to increase its utility in different domains. DT is generating a virtual imitation of a physical object that mimics the status of an event by changing the information in real time. |
| 158 | 10.3390/fi15070223 | Avanzato | 2023 | Digital replicas of patients' hearts, thus assisting in clinical decision making and testing of new therapies. |
| 160 | 36908269 | Susilo | 2023 | A virtual representation of a mosunetuzumab-treated clinical patient that integrates patient-specific clinical data (i.e., pharmacokinetics [PKs], tumor size, and biomarker data) alongside other in vitro/in vivo data within the established mosunetuzumab QSP model |
| 165 | 37363928 | Strocchi | 2023 | A patient-specific anatomical heart model, or digital twin, |
| 166 | 37160583 | Shu | 2023 | Digital twins are virtual counterparts of real-world processes, modeling dynamics and properties in real-time [1]. Receiving continuous measurements from sensor-rich environments, digital twins can conversely provide computational feedback, which is dificult to obtain otherwise. |

**Organs and systems represented by the claimed digital twins.**

| Organ | n = 48 |
|---|---|
| Heart | 15 (31%) |
| Bones and joints | 10 (21%) |
| Lung | 6 (12%) |
| Arteries | 5 (10%) |
| Craniofacial structure – Skull - Teeth | 3 (6%) |
| Multiple organs | 3 (6%) |
| Gastrointestinal tract | 2 (4%) |
| Brain | 1 (2%) |
| Breast | 1 (2%) |
| Liver | 1 (2%) |
| Skin | 1 (2%) |
| **System** | **n = 11** |
| Endocrine system | 4 (36%) |
| Immune system | 3 (27%) |
| Haematological system | 1 (9%) |
| Lymphatic system | 1 (9%) |
| Neuromuscular system | 1 (9%) |
| Reproductive system | 1 (9%) |

**Medical disciplines represented.**

| Medical disciplines | Total<br>n = 80 |
|---|---|
| Cardiology | 16 (20 %) |
| Oncology | 10 (12.5%) |
| • General | 3 |
| • Breast | 3 |
| • Haematological (lymphoma and leukaemia) | 2 |
| • Colorectal | 1 |
| • Prostate | 1 |
| Orthopaedics | 9 (11%) |
| Intensive care | 5 (6.2%) |
| Diabetology | 4 (5.0%) |
| Neurology | 4 (5.0%) |
| Vascular medicine | 4 (5.0%) |
| Immunology | 3 (3.8%) |
| Pharmacology | 3 (3.8%) |
| Radiology | 3 (3.8%) |
| Dentistry/Orthodontics | 3 (3.8%) |
| Surgery | 3 (3.8%) |
| • General | 1 |
| • Lung | 1 |
| • Neurosurgery | 1 |
| Endocrinology | 2 (2.5%) |
| Gastroenterology | 2 (2.5%) |
| Geriatrics | 2 (2.5%) |
| Obstetrics and gynecology | 2 (2.5%) |
| Dermatology | 1 (1.3%) |
| Hepatology | 1 (1.3%) |
| Other (not classifiable) | 3 (3.8%) |
| • Emotion recognition | 1 |
| • Sports medicine | 1 |
| • Well-being | 1 |

**Data used to develop and/or run the 80 claimed digital twins.**

| Data used to develop and/or run the claimed digital twins | Total<br>n = 80 |
|---|---|
| **Data from clinical notes** | **15 (19%)** |
| **Laboratory data** | **13 (16%)** |
| **Imaging data** | **35 (44%)** |
| • CT | 13/35 (37%) |
| • MRI | 9/35 (26%) |
| • CT + MRI | 4/35 (11%) |
| • X-rays | 3/35 (8.6%) |
| • US | 2/35 (5.7%) |
| • CT + US | 1/35 (2.9%) |
| • OCT | 1/35 (2.9%) |
| • OCT + Angiography | 1/35 (2.9%) |
| • Not specified | 1/35 (2.9%) |
| **Wearable device data** | **12 (15%)** |
| • Continuous glucose monitoring | 4/12 (33.3%) |
| • Combination of actimeters, mobile apps, ± other wearables (smartscales, continuous glucose monitoring) | 2/12 (16.6%) |
| • Connected sleeve | 1/12 (9.1%) |
| • Smartwatches | 1/12 (9.1%) |
| • Inertial measurement units | 1/12 (9.1%) |
| • Smart insole | 1/12 (9.1%) |
| • Actimeters | 1/12 (9.1%) |
| **Other data sources*** | **32 (40%)** |
| • ECG | 5/32 (15.6%) |
| • Invasive pressure measurements | 4/32 (12.5%) |
| • Optical tracking and motion capture | 4/32 (12.5%) |
| • Treatment data | 4/32 (12.5%) |
| • Ventilators parameters | 4/32 (12.5%) |
| • Diet data | 3/32 (9.4%) |
| • Vital signs | 3/32 (9.4%) |
| • Electro-anatomical mapping data | 2/32 (6.3%) |
| • Video data | 2/32 (6.3%) |
| • Cardiotocography recordings | 1/32 (3.1%) |
| • Endoscopic evaluation | 1/32 (3.1%) |
| • Genotype | 1/32 (3.1%) |
| • Impedance tomography data | 1/32 (3.1%) |
| • Lung function test | 1/32 (3.1%) |
| • Pathological data | 1/32 (3.1%) |
| • Patient report of exertion | 1/32 (3.1%) |
| • Performance metrics evaluated by coaches | 1/32 (3.1%) |
| • Simulated data | 1/32 (3.1%) |
| • Wound pictures | 1/32 (3.1%) |

*The sum of each data sources exceeds 32 because one claimed digital twin could include several other data sources CT: computed tomography, MRI: Magnetic Resonance Imaging, OCT: Optical coherence tomography, US: ultrasound.

### Update Frequency

| Frequency of input of new data into the digital twin | Total<br>n = 20 |
|---|---|
| Real-time | 14 (70%) |
| Hour | 2 (10%) |
| Day | 4 (20%) |
| **Intended output frequency for the dynamic digital twin** | |
| Real-time | 7 (35%) |
| Hour | 2 (10%) |
| Day | 7 (35%) |
| Not specified | 4 (20%) |

**Unsupervised classification of claimed digital twins**

| Characteristic | Cluster 1<br>n = 43 | Cluster 2<br>n = 22 | Cluster 3<br>n = 15 | p-value |
|---|---|---|---|---|
| Multimodal data | | | | <0.001 |
| • yes | 27 (63%) | 12 (55%) | 0 (0%) | |
| • no | 16 (37%) | 10 (45%) | 13 (87%) | |
| • unclear | 0 (0%) | 0 (0%) | 2 (13%) | |
| Multiscale (yes) | 4 (9.3%) | 0 (0%) | 5 (33%) | 0.007 |
| Multiphysics (yes) | 5 (12%) | 0 (0%) | 3 (20%) | 0.094 |
| Type of model (organ, system or other) | | | | <0.001 |
| • Organ | 36 (84%) | 8 (36%) | 4 (27%) | |
| • System | 6 (14%) | 1 (4.5%) | 4 (27%) | |
| • Other | 1 (2.3%) | 13 (59%) | 7 (47%) | |
| Type of model (anatomical and/or physiological) | | | | <0.001 |
| • Anatomical | 7 (16%) | 3 (14%) | 0 (0%) | |
| • Physiological | 13 (30%) | 1 (4.5%) | 8 (53%) | |
| • Both | 23 (53%) | 0 (0%) | 5 (33%) | |
| • Other | 0 (0%) | 18 (82%) | 2 (13%) | |
| Approach | | | | <0.001 |
| • Data-driven | 1 (2.3%) | 20 (91%) | 1 (6.7%) | |
| • Mechanistic | 31 (72%) | 0 (0%) | 9 (60%) | |
| • Both | 10 (23%) | 2 (9.1%) | 5 (33%) | |
| • Other | 1 (2.3%) | 0 (0%) | 0 (0%) | |
| Model for simulation (versus simulation itself) | 42 (98%) | 2 (9.1%) | 12 (80%) | <0.001 |
| Type of CDT (static or dynamic) | | | | <0.001 |
| • Static | 36 (84%) | 8 (36%) | 0 (0%) | |
| • Dynamic | 6 (14%) | 13 (59%) | 1 (6.7%) | |
| • Not classifiable (not patient specific) | 1 (2.3%) | 1 (4.5%) | 14 (93%) | |
| Type of data exchange | | | | <0.001 |
| • One-way | 39 (91%) | 16 (73%) | 3 (20%) | |
| • Two-way | 3 (7.0%) | 5 (23%) | 1 (6.7%) | |
| • No | 1 (2.3%) | 1 (4.5%) | 11 (73%) | |
| Patient-specific CDT (yes) | 42 (98%) | 21 (95%) | 1 (6.7%) | <0.001 |
| Data analytics (yes) | 41 (95%) | 22 (100%) | 15 (100%) | 0.7 |
| Advanced visualisation (yes) | 26 (60%) | 0 (0%) | 3 (20%) | <0.001 |
| Objective: simulation (yes) | 37 (86%) | 1 (4.5%) | 14 (93%) | <0.001 |
| Objective: prediction (yes) | 34 (79%) | 18 (82%) | 9 (60%) | 0.3 |
| Objective: monitoring (yes) | 4 (9.3%) | 7 (32%) | 0 (0%) | 0.014 |
| Objective: visualisation (yes) | 10 (23%) | 0 (0%) | 0 (0%) | 0.007 |
| Objective: generation (yes) | 3 (7.0%) | 0 (0%) | 5 (33%) | 0.004 |

Please refer to appendix pp 5-6 for more explanations on the variables used.
